# Developing a natural language processing system using transformer-based models for adverse drug event detection in electronic health records

**DOI:** 10.1101/2024.07.09.24310100

**Authors:** Jingyuan Wu, Xiaodi Ruan, Elizabeth McNeer, Katelyn M. Rossow, Leena Choi

## Abstract

**Objective:** To develop a transformer-based natural language processing (NLP) system for detecting adverse drug events (ADEs) from clinical notes in electronic health records (EHRs).

**Materials and Methods:** We fine-tuned BERT Short-Formers and Clinical-Longformer using the processed dataset of the 2018 National NLP Clinical Challenges (n2c2) shared task Track 2. We investigated two data processing methods, window-based and split-based approaches, to find an optimal processing method. We evaluated the generalization capabilities on a dataset extracted from Vanderbilt University Medical Center (VUMC) EHRs.

**Results:** On the n2c2 dataset, the best average macro F-scores of 0.832 and 0.868 were achieved using a 15-word window with PubMedBERT and a 10-chunk split with Clinical-Longformer. On the VUMC dataset, the best average macro F-scores of 0.720 and 0.786 were achieved using a 4-chunk split with PubMedBERT and Clinical-Longformer.

**Discussion:** Our study provided a comparative analysis of data processing methods. The fine-tuned transformer models showed good performance for ADE-related tasks. Especially, Clinical-Longformer model with split-based approach had a great potential for practical implementation of ADE detection. While the token limit was crucial, the chunk size also significantly influenced model performance, even when the text length was within the token limit.

**Conclusion:** We provided guidance on model development, including data processing methods for ADE detection from clinical notes using transformer-based models. Our results on two datasets indicated that data processing methods and models should be carefully selected based on the type of clinical notes and the allocation trade-offs of human and computational power in annotation and model fine-tuning.

## BACKGROUND AND SIGNIFICANCE

Adverse drug events (ADEs) refer to any physical or psychological injuries and unexpected events caused by medication use,[1] and can be significant hospital complications impacting patient experience and costs.[2] It is challenging to detect ADEs at an early stage due to ambiguous and incomprehensive information about actions, symptoms and medications. Thus, information extracted from electronic health records (EHRs), including diagnoses, prescriber notes, laboratory tests and results, becomes important in supporting early treatment and primary and secondary prevention.

However, extracting relevant information from unstructured clinical notes is challenging. Over the past few decades, several approaches have been used including rule-based, machine learning-based, deep learning-based, and contextualized language model-based approaches.[3] More recently, contextualized language models are widely applied in natural language processing (NLP). These models are pre-trained on large corpora and have a good understanding of the contextual patterns in the relevant domain, which promotes their application in downstream tasks. The Bidirectional Encoder Representations from Transformer (BERT) model,[4,5] one of the most widely used contextualized models, was first introduced in 2018, which employs a transformer architecture to pre-train a deep neural network on a large corpus of unlabeled text data, using an unsupervised approach. Since BERT has continuously evolved, BERT-based NLP systems have shown great promise for ADE-related tasks, such as relationship extraction and text classification.[3,6] Fan *et al.*[7] conducted a comparative analysis between BERT-based models, standard deep learning models, and current state-of-the-art models for ADE detection and extraction, and demonstrated that a BERT-based model achieved new state-of-the-art results. Hussain *et al.*[8] proposed an end-to-end system for adverse drug relation detection by fine-tuning BERT, showing good performance on Twitter and PubMed datasets. The introduction of SpanBERT[9] architecture to ADE extraction task outperformed competing models[10] on Social Media Mining for Health (SMM4H) and CSIRO Adverse Drug Event Corpus (CADEC).[11,12] Narayanan *et al.*[13] evaluated various biomedical contextual embeddings and models using the 2018 National NLP Clinical Challenges (n2c2) shared task Track 2 data on ADEs and Medication Extraction,[14] demonstrating the importance of BERT structure. In recent work on adverse event (AE) detection, Chopard *et al.*[15] confirmed the feasibility of automating coding of AEs described in the narrative section of serious AE report forms. Furthermore, Silverman *et al.*[16] demonstrated that large language models (LLMs) such as University of California – San Francisco (UCSF)-BERT achieved higher accuracy in serious AE detection from clinical notes compared with previous methods. A recent review[17] on machine learning and deep learning approaches in ADE extraction for benchmark datasets, including n2c2,[14] highlighted BERT’s superior model performance in end-to-end tasks.

Despite the good performance of BERT model and its variants (we call them BERT Short-Formers hereafter as opposed to Longformer,[18] which we will discuss below), one of their limitations is that long text may be truncated due to the token limit. To overcome this issue, Longformer,[18] a BERT variant that can handle long sequences of text, was developed. The key innovation of Longformer is its attention mechanism, which considers window-based local-context self-attention and global attention to achieve powerful contextual and sequence representations.

Our work began with exploring an effective data processing method for extracting relevant information from clinical notes in EHRs for transformer-based models, ultimately aiming to develop an NLP system to detect ADEs in clinical notes. To achieve this goal, we systematically investigated two data processing methods, called window-based and split-based approaches, to generate model inputs for fine-tuned BERT Short-Formers (BERT-base-uncased,[5] Biomed_Roberta,[19] Bio_ClinicalBERT,[20] BioBERT,[21] PubMedBERT,[22] SpanBERT[9]) and Clinical-Longformer[23,24] on the n2c2 data. We then applied optimal processing and modeling methods to a dataset extracted from Vanderbilt University Medical Center (VUMC) EHRs to further evaluate generalization capabilities. The two datasets contain very different types of clinical notes: the n2c2 data has fewer but longer notes with more ADEs associated with various drugs, while the VUMC data has more but shorter notes with homogeneous ADEs related to two drugs of interest. By undertaking these different approaches on two distinct datasets, our work provided guidance for identifying ADEs from free text using transformer-based models.

## MATERIALS AND METHODS

### The Data

#### The n2c2 data

The n2c2 data was from the 2018 n2c2 shared task Track 2.[14,25–29] For this study, we used existing gold standard labels focusing on the drug names, ADEs, and their relationships. 505 clinical notes were divided into two sets: 303 notes in the training set, 78.20% containing ADE relations, and 202 notes in the test set, 76.73% containing ADE relations.

#### The VUMC data

The VUMC data was extracted from EHRs at VUMC and was previously used in another study[30] that included a cohort of pediatric patients with ADEs related to two drugs of interest (citalopram/Celexa, escitalopram/Lexapro). As the gold standard labels were not available, we annotated the gold standards as described below. Our final cohort included 112 patients with a total of 1,541 notes that mentioned a drug of interest, split into: 1,109 notes in the training set, 12.17% containing ADE relations, and 432 notes in the test set, 8.33% containing ADE relations.

### Annotation of the VUMC Data

To create gold standard labels for the VUMC data, we used the brat annotation tool (BRAT)[31] to manually annotate AEs associated with drugs in clinical notes. Before labelling, we formulated an initial set of annotation labels and corresponding annotation guidelines. These guidelines and the annotation labels were updated during an iterative process where a few notes were annotated, discussed, and reannotated within a small group of researchers.

Specifically, we started with detailed labels to reflect clinical context: AEpositive, AEnegative, AEpositiveNoJustification, AEconditional, and NoResponse. Drug names were labeled with AEpositive if they were associated with an ADE, AEnegative if they were not associated with an ADE, AEpositiveNoJustification if they were associated with a change in dose due to an ADE but did not provide its justification, AEconditional if they were associated with a possible change in dose due to an ADE if certain conditions were met (e.g., if symptoms such as weight loss worsened while on the medication), and NoResponse if they were associated with a change in dose due to non-response to drug (i.e., drug is not working), but not due to an ADE. After completion of the annotation, we randomly selected a subset of 300 notes to validate the annotation by two independent annotators and evaluated agreement using a Kappa statistic. The Kappa statistic was 0.820. Discrepancies between the annotators were discussed as a group and a final decision was made about what the gold standard should be.

From these initial labels, for our ADE classification task, the gold standard label for “ADE positive” was created by combining AEpositive, AEpositiveNoJustification, and AEconditional labels, while the label for “ADE negative” was created by combining AEnegative and NoResponse labels.

### Data Processing

Pre-trained transformer models can take a limited length of free text, typically 512 tokens, whereas the Longformer can take up to 4,096 tokens. As clinical notes are typically unstructured and long, the text should be processed before modeling. We investigated several processing methods, eventually focusing on the following two.

#### Window-based approach

When drug names of interest are already annotated or can be located easily, this approach can be useful to effectively capture the contextual details surrounding drug names. It extracts a specific number of words before and after the drug name, called *window size*, by leveraging the definition of words using spaces; for example, 20 words before and after vancomycin, a drug of interest for the n2c2 dataset. The window sizes we investigated include 10, 15, 20, 50, and 100 words. If the number of words before or after the drug name was insufficient, we would extract words up to the beginning or end of each note.

#### Split-based approach

We explored a simpler alternative when drug names are not yet annotated, requiring substantial effort to locate. This approach exclusively partitions clinical notes into pre-defined *chunk sizes* such as 2, 4, 10, and 20 chunks. After investigating the effects of splitting notes at both word and sentence levels, we found that splitting at sentence level generally performed better (see Supplemental Material) as it enhanced comprehension by including complete context. Therefore, all results we reported were based on sentence-level splits. We first used the Natural Language Toolkit (NLTK) tokenizer[32] to tokenize the text to sentences, subsequently, dividing the sentences into pre-defined chunk size. If the token length of a particular chunk exceeded a pre-defined token limit (e.g., 4,096 for Longformer), we implemented a further division of that chunk into two smaller chunks at the word level. This additional step ensured that the chunks remained within the desired token limit, preventing truncation of context. After this, no further splitting would be required as most chunks were within the token limit, while a very few chunks might still exceed the token limit due to missing punctuation in some parts of the clinical notes, especially at the beginning or end.

For both approaches, newline characters were replaced by spaces for easier processing. A schematic diagram of data processing steps and examples of different processing methods are shown in Figure 1.

**Figure 1.**
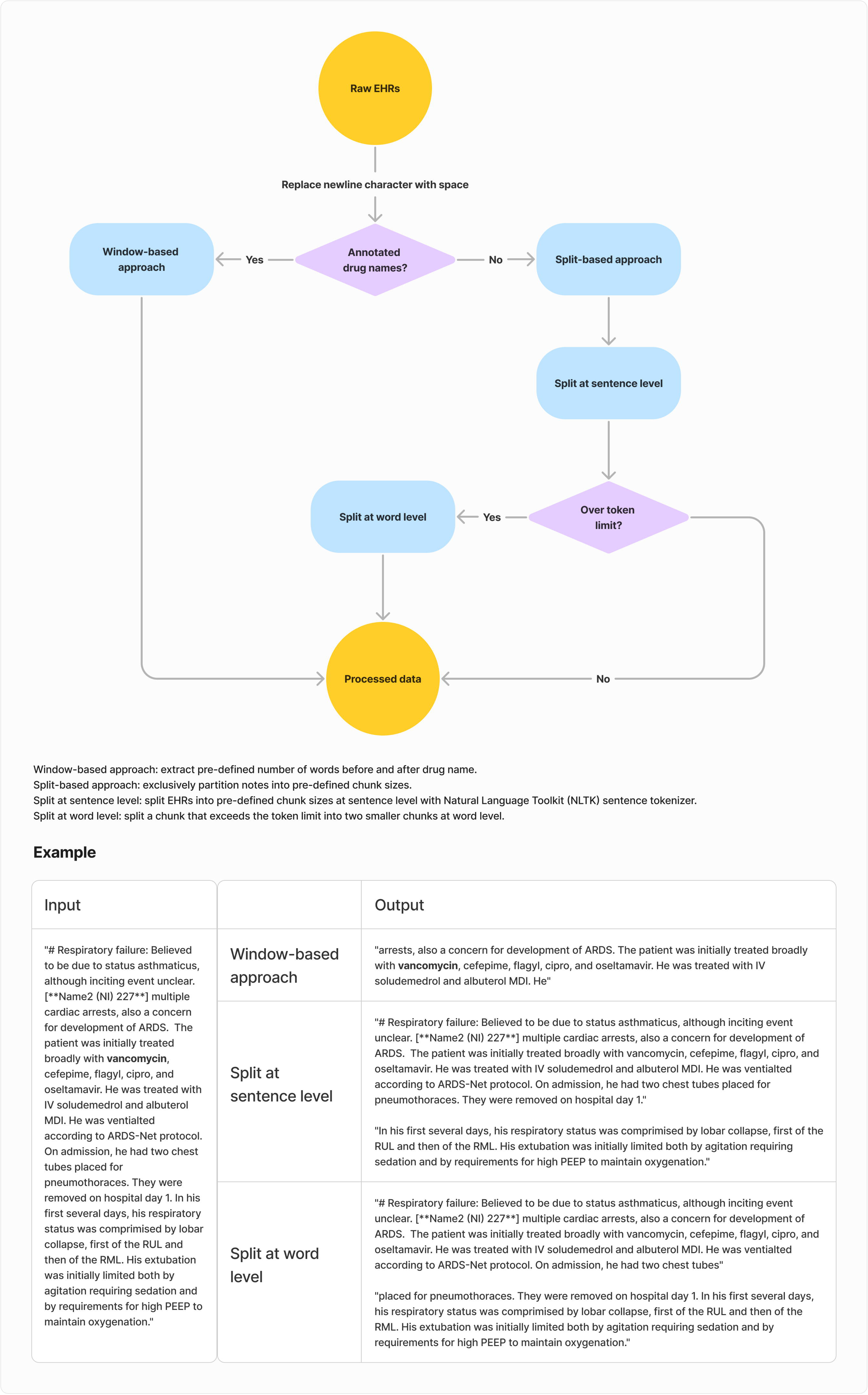
Schematic diagram of data processing steps and examples of data processing methods.

### Model Fine-Tuning

#### BERT Short-Formers

For fine-tuning BERT Short-Formers, we tried different batch sizes, epochs, learning rates and early stop mechanisms, but none of these showed potential to significantly improve model performance with our computing resources. Therefore, we used batch size per device of 16 and 5 GPUs. All other default settings were retained (e.g., epoch of 3, AdamW optimizer with a learning rate of 5e-5, no warmup ratio or early stopping applied).

#### Clinical-Longformer

For fine-tuning the Clinical-Longformer model, we employed an adaptive attention window by considering the distribution of sequence lengths of the split chunks. Compared to the n2c2 dataset, the text length of the VUMC dataset is much shorter. Thus, when fitting the VUMC dataset, we set a custom warmup ratio to let the learning rate start at a relatively low value and then gradually increase over the training steps. This allowed the model to explore a wider range of solutions at the early stage and prevented it from getting stuck in suboptimal solutions by providing a controlled way to increase the learning rate. We used default attention window of 512 and epoch of 3 for the n2c2 dataset, and attention window of 64, epoch of 10, early stopping patience of 3, and warmup ratio of 0.3 for the VUMC dataset. For both datasets, batch size per device of 2 and 8 GPUs were used, and all others were default settings. All work was done through Hugging Face.[33]

### Evaluation Metrics

Extremely imbalanced datasets are common in ADE studies because ADEs are often rare events and hence the frequency of positive ADEs is much smaller than that of negative. On the other hand, since preventing false negatives is as important as false positives, an F-score is used as a model performance metric that considers both precision and recall. The micro F-score measures the model predictions with a weighted mean, while the macro F-score computes an unweighted mean. Even though we found that the model performance evaluated using micro F-score was consistently much higher, we used the macro F-score as our main evaluation metric (supplemented by precision and recall), as we want to measure the predictions of the two classes equally, regardless of the sample size of the two classes.

With the actual values in the data and the model predicted values, the final classification results generated by a classifier are true positive (TP), true negative (TN), false positive (FP), and false negative (FN). The equation of macro F-score for binary classification is as follows:

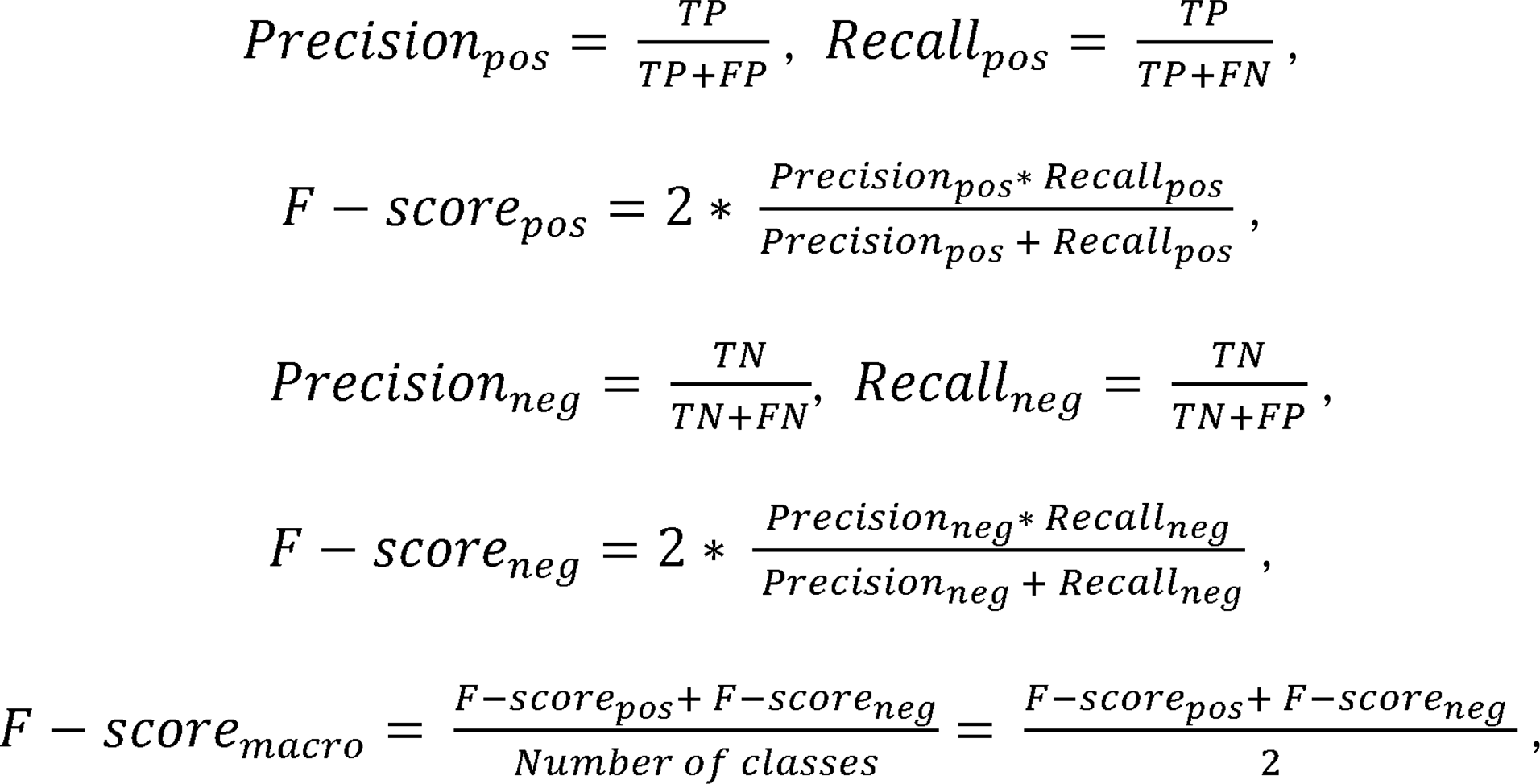

where pos and neg denote positive and negative, respectively. We evaluated the performance of each model by the average of three trials with different train-test split seeds for validation. Due to the inconsistency of Clinical-Longformer when applying non-default attention windows, we conducted three trials each with three different train-test split seeds, leading to a total of nine trials instead of three for validation.

## RESULTS

Table 1 presents the results for the fine-tuned Bert-base-uncased model on the n2c2 dataset using varying window sizes in the window-based data processing. Window sizes of 10 to 20 words provided a good performance with 15-word yielding the best result with F-score of 0.805. This confirmed our initial conjecture that a 15-word window around the drug name would provide sufficient information about ADEs associated with the drug. Our exploratory analysis regarding the distance between drug names and ADEs found that a 15-word window covered approximately 80% of ADEs. Wider window sizes yielded poorer performance, which could be attributed to the potential challenges posed by longer texts with more complex contextual information (e.g., medical history and prescription). While wider windows appear to be beneficial in capturing a wider range of context, they do not guarantee better model performance. On the other hand, if the window size is too small, the excerpt may not include ADEs, lowering the recall, while we would like to detect as many positive cases as possible.

**Table 1.** Effects of Window Size on Model Performance of Bert-base-uncased using the n2c2 Dataset.

| Window Size | Average of Precision | Average of Recall | Average of F-score |
| --- | --- | --- | --- |
| 10-word | 0.845 | 0.775 | 0.805 |
| 15-word | 0.834 | **0.784** | **0.805** |
| 20-word | **0.855** | 0.757 | 0.796 |
| 50-word | 0.801 | 0.698 | 0.736 |
| 100-word | 0.755 | 0.648 | 0.683 |

To leverage domain-specific knowledge, we fine-tuned several BERT Short-Formers that were pre-trained on biomedical corpora. Using the processing method with the window size of 15 words selected as the best window size with Bert-base-uncased model, we assessed their model performance improvement using the n2c2 dataset (Table 2). PubMedBERT provided the best performance with F-score of 0.832, which indicates that the BERT model pre-trained on a biomedical corpus was effective in capturing domain-specific information and enhancing the overall performance. Additionally, the Clinical-Longformer achieved the best precision, further highlighting the benefits of leveraging a BERT model pre-trained on clinical text. Therefore, in the following tasks, Clinical-Longformer was used instead of Longformer for better classification purpose.

**Table 2.**
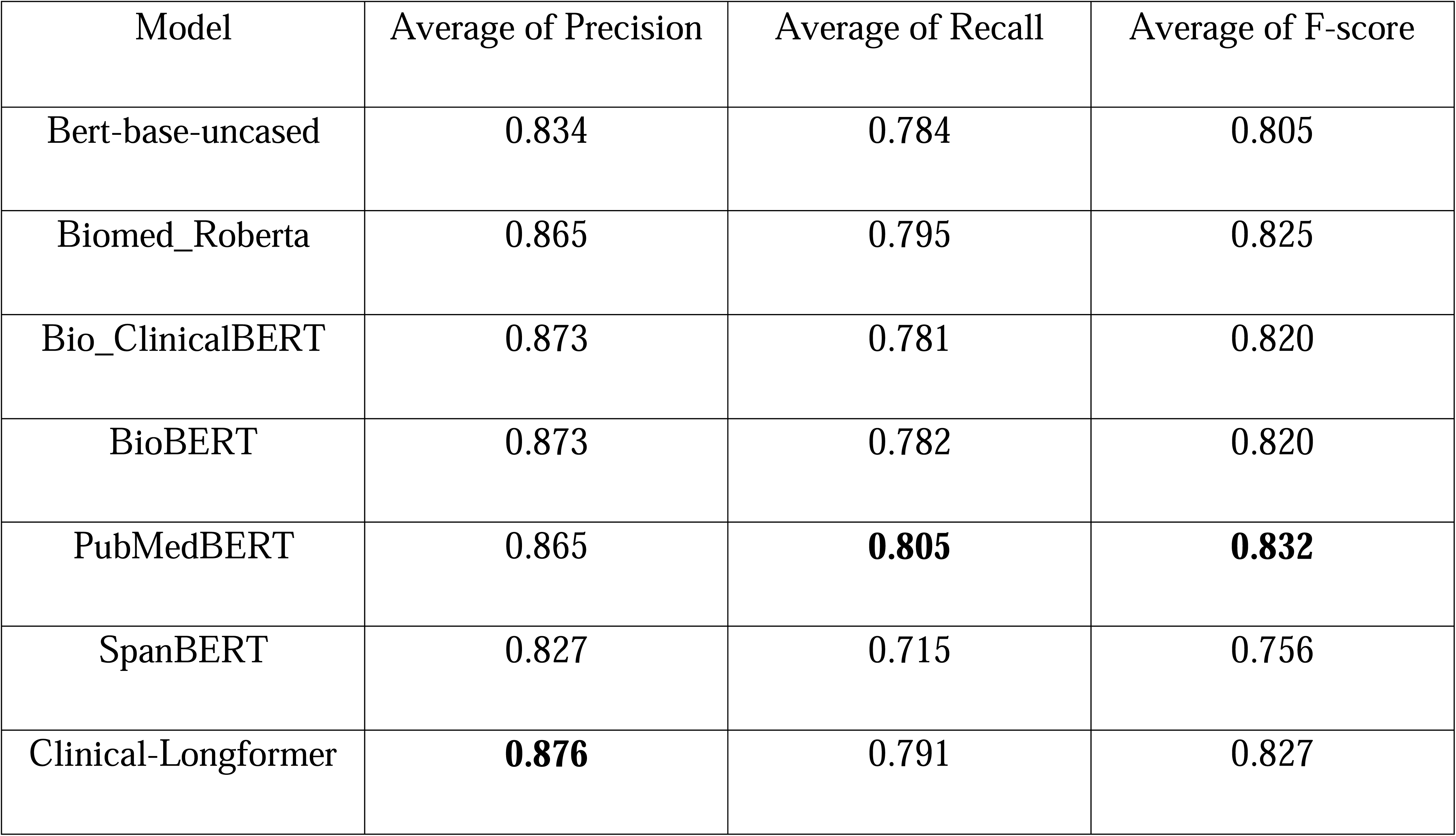
Model Comparison with Window Size of 15 Words using the n2c2 Dataset.

| Model | Average of Precision | Average of Recall | Average of F-score |
| --- | --- | --- | --- |
| Bert-base-uncased | 0.834 | 0.784 | 0.805 |
| Biomed_Roberta | 0.865 | 0.795 | 0.825 |
| Bio_ClinicalBERT | 0.873 | 0.781 | 0.820 |
| BioBERT | 0.873 | 0.782 | 0.820 |
| PubMedBERT | 0.865 | **0.805** | **0.832** |
| SpanBERT | 0.827 | 0.715 | 0.756 |
| Clinical-Longformer | **0.876** | 0.791 | 0.827 |

The results of text partitioning on the BERT Short-Formers and Clinical-Longformer using the n2c2 dataset are presented in Table 3. It revealed that the number of chunks (i.e., chunk size) played a critical role on the model performance as it determines the proportion of text truncated due to token limit. Overall, Clinical-Longformer achieved good performance on most of the chunk sizes, where almost all chunks are within its token limit: 10-chunk split provided the best F-score of 0.868, followed by 20 and 4 chunks with F-scores of 0.850 and 0.841, respectively. We also conducted a comparative analysis of the performance between 10 and 20 chunks across two BERT Short-Formers [Bert-base-uncased (baseline), PubMedBERT (best performer in Table 2)] and the Clinical-Longformer using the n2c2 dataset. For this comparison, chunk sizes of 10 and 20 were selected, as most chunks were within the token limit of BERT Short-Formers, considering the 95th percentile word count was about 1,368 in the n2c2 training set. The results of 2 and 4 chunks were not provided for the BERT Short-Formers as most chunks would exceed the token limit. For Bert-base-uncased, PubMedBERT, and Clinical-Longformer, we obtained F-scores of 0.715, 0.794, and 0.868 with 10 chunks, while 0.761, 0.823, and 0.850 with 20 chunks, respectively. The results between these two chunk sizes emphasize the importance of selecting the appropriate partitioning strategy to ensure context integrity for optimal model performance.

**Table 3.** Effects of Split Chunk Size on Model Performance using the n2c2 Dataset.

| Model | Chunk Size | Average of Precision | Average of Recall | Average of F-score | Proportion over Token Limit |
| --- | --- | --- | --- | --- | --- |
| Bert-base-uncased | 2 chunks | - | - | - | - |
|  | 4 chunks | - | - | - | - |
|  | 10 chunks | 0.752 | 0.699 | 0.715 | 0.070 |
|  | 20 chunks | 0.806 | 0.730 | 0.761 | 0.026 |
| PubMedBERT | 2 chunks | - | - | - | - |
|  | 4 chunks | - | - | - | - |
|  | 10 chunks | 0.815 | 0.778 | 0.794 | 0.057 |
|  | 20 chunks | 0.838 | 0.804 | 0.823 | 0.022 |
| Clinical-Longformer | 2 chunks | 0.683 | 0.674 | 0.674 | 0.002 |
|  | 4 chunks | 0.837 | 0.850 | 0.841 | 0.000 |
|  | 10 chunks | **0.865** | **0.872** | **0.868** | 0.000 |
|  | 20 chunks | 0.850 | 0.850 | 0.850 | 0.000 |

Table 4 shows the performance for two selected BERT Short-Formers and Clinical-Longformer for classifying ADEs in the VUMC dataset. As clinical notes in the VUMC dataset are relatively short, we processed the data using split chunks size of 1 (i.e., entire note, so no split), 2, 4 and 6 chunks. The results of 1-chunk with BERT Short-Formers are not provided because they didn’t converge due to a high proportion of chunks exceeding the token limit. Although the VUMC data are mostly within the token limit of the Longformer, the 2-chunk and 4-chunk still performed better than 1-chunk. This may be because a text with appropriate length could provide better context to capture ADEs more precisely. Similar findings were obtained with Bert-base-uncased and PubMedBERT. Furthermore, Clinical-Longformer significantly outperformed BERT Short-Formers as it can handle eight times longer tokens to ensure more complete context. However, 6-chunk did not improve performance because further splitting may increase the chance that ADE and drug name are in separate chunks. A higher number of chunks fragments the entire text into more pieces, posing a challenge for the model to capture the comprehensive context of the text. The deterioration in model performance with an increasing number of chunks is also likely attributable to the trade-off between model complexity and fit. While a higher number of chunks simplifies text input, it concurrently elevates model complexity as the model is compelled to handle more input segments.

**Table 4.** Effects of Split Chunk Size on Model Performance using the VUMC Dataset.

| Model | Chunk Size | Average of Precision | Average of Recall | Average of F-score | Proportion over Token Limit |
| --- | --- | --- | --- | --- | --- |
| Bert-base-uncased | 1 chunk | - | - | - | 0.444 |
|  | 2 chunks | 0.654 | 0.639 | 0.644 | 0.194 |
|  | 4 chunks | 0.661 | 0.659 | 0.657 | 0.067 |
|  | 6 chunks | 0.689 | 0.736 | 0.708 | 0.042 |
| PubMedBERT | 1 chunk | - | - | - | 0.436 |
|  | 2 chunks | 0.677 | 0.651 | 0.651 | 0.177 |
|  | 4 chunks | 0.727 | 0.731 | 0.720 | 0.059 |
|  | 6 chunks | 0.728 | 0.716 | 0.712 | 0.037 |
| Clinical-Longformer | 1 chunk | 0.713 | 0.701 | 0.704 | 0.012 |
|  | 2 chunks | 0.742 | 0.788 | 0.762 | 0.000 |
|  | 4 chunks | **0.774** | 0.801 | **0.786** | 0.000 |
|  | 6 chunks | 0.725 | **0.813** | 0.758 | 0.000 |

## DISCUSSION

We fine-tuned various pre-trained transformer-based models for detecting ADEs from clinical notes on two different datasets, each with different structure and content. Among BERT Short-Formers, the PubMedBERT model performed best with F-score of 0.832, while the Clinical-Longformer model outperformed BERT Short-Formers with F-score of 0.868 on the n2c2 dataset. On the VUMC dataset, Clinical-Longformer model consistently outperformed BERT Short-Formers with best F-score of 0.786. To the best of our knowledge, this is the first study to fine-tune the Longformer model for ADE detection.

One of the major limitations of transformer models is that they cannot take long text that exceeds the token limit. Thus, we explored several data processing approaches to find the optimal method for extracting key information from EHRs for ADE detection. The window-based approach can efficiently extract most relevant excerpts surrounding the drug of interest. We found that a range of 10 to 20 words window size provided similarly good performance with maximum performance at 15 words. Our results suggest the importance of selecting an optimal window size to balance between context and complexity. As this approach requires annotated drug names, we aimed to find a simpler alternative, called the split-based approach. This approach can be useful for real-world implementation as clinical notes typically lack annotated drug names. As the number of partitions needs to be pre-determined, we compared different chunk sizes and their impact on model performance. We found that the number of chunks impacted model performance, even if the note length was within the token limit. As the optimal chunk size depends on the note styles and context, we recommend assessing different chunk sizes to determine an optimal one for any given dataset or performing sensitivity analysis.

Additionally, we compared the model performance between the processing methods (window vs. split) with the same model as well as between BERT Short-Formers and Longformer with the same processing method. PubMedBERT performed slightly better with the 15-word window compared to 20-chunk split. This may be because the window-based approach can build ideally more precise context surrounding drug name. In contrast, Clinical-Longformer performed slightly better with the split-based approach. This may be because the window-based approach doesn’t take full advantage of Longformer for handling long texts. In addition, PubMedBERT and Clinical-Longformer are trained in different ways and on different corpora, which can lead to different model characteristics. These comparisons shed light on the differences between models and their ability for given tasks.

There are a few previous works on the n2c2 dataset for the ADE-Drug relation, focusing on BERT Short-Formers. Most of our fine-tuned BERT Short-Formers performed better than the best models reported by Wei *et al.* (F-score of 0.80 on BERT-large-uncased and 0.81 on MIMIC BERT).[5,6,34] Mahendran *et al.*[3] reported the best F-scores of 0.97 and 0.97 on Bert-base-uncased[5] and BioBERT,[21] respectively, but it is not clear whether these scores are micro or macro F-scores for ADE-Drug relation. Our reimplementation of these models, however, suggested that micro F-score was likely to be utilized in these papers. Our micro F-scores were in a similar range (e.g., 0.96 on 15-word window Bert-base-uncased), but we did not use micro F-score as we found that it was not able to differentiate model performance across the models and the methods since all scores are very high, and hence we chose macro F-score as a more appropriate metric for evaluation.

When modeling with the VUMC dataset, we found Clinical-Longformer showed less stability and slower convergence than BERT Short-Formers when using non-default attention windows. Despite its ability to handle longer text, Clinical-Longformer exhibited erratic behavior, including sudden performance drops, quick overfitting, and occasional failure to converge. However, even when overfit, transformer models could still offer valuable insights for downstream tasks. Furthermore, our study specifically focuses on AEs associated with two drugs, citalopram (brand name Celexa) and escitalopram (brand name Lexapro), using data obtained from a previous study investigating AEs for these drugs.

Despite our efforts, our work has some limitations. Firstly, due to limited GPU resources, we were constrained to use a small batch size per device during Clinical-Longformer training. This restriction may not have fully explored Longformer’s capabilities for these tasks, and further investigations with larger batch sizes could provide more insights. Secondly, our evaluation was primarily focused on the n2c2 dataset and one dataset of our own, which may provide limited diversity in the data types and clinical note styles. This limitation could impact the generalizability of our findings to other datasets. Thirdly, the presence of annotation bias should be acknowledged. Differences among annotators, training processes, and potential human errors may have introduced bias to the annotated data. Lastly, while our methods were tested on BERT Short-Formers and Clinical-Longformer, other transformer models may offer untapped potential for these tasks.

## CONCLUSION

In this study, we presented transformer-based models for detecting ADEs as well as various data processing methods for their impact on the model performance. Window-based processing approaches rely heavily on annotations, which is labor-intensive, but can build precise context when the distances between drug names and their ADEs in the dataset are relatively consistent. Split-based processing approaches can easily split long texts into smaller chunks transformer models can handle and does not require annotation for Drug-ADE relations. However, chunk size determines the proportion of chunks that exceed the token limit, which affects the final classification task, and determining the optimal chunk size can be challenging. The Longformer model requires more fine-tuning work, including but not limited to memory, time and hyperparameter settings, while for BERT Short-Formers, token limit is the major challenge.

Although our results are promising, the limitations highlight areas for further research and improvements to enhance the applicability and generalizability of transformer-based NLPs for ADE detection in real-world healthcare systems.

For future work, we aim to extend our findings to a variety of datasets, allowing for more comprehensive validation across clinical scenarios. To enhance the extraction of ADE-related information from EHRs, we can consider additional aspects such as symptoms, actions, and dosage changes. This enriched information can contribute to a more comprehensive understanding of ADE occurrences, which can support better clinical decision-making and patient care. Ultimately, our ongoing research will continue to drive innovation and advancements in the field of ADE detection, contributing to the development of robust and practical NLPs to improve patient safety and healthcare outcomes.

## ACKNOWLEDGEMENT

We thank Sara L. Van Driest and Ida Aka for providing the VUMC dataset and clinical context, which helped the annotation process.

## FUNDING

This work is supported by NIH/NIGMS (R01-GM124109) and NIH/NICHD P50 HD106446-01S1.

## AUTHOR CONTRIBUTIONS

LC conceived the research and managed the project. JW developed the models and performed analyses. JW, XR, and LC drafted the manuscript. EM, JW, and XR annotated gold standards for the VUMC data. KMR collected the VUMC dataset. All authors reviewed and edited the final manuscript.

## CONFLICTS OF INTEREST

None declared.

## DATA AVAILABILITY STATEMENT

The n2c2 data is open source at https://n2c2.dbmi.hms.harvard.edu/data-sets.

## Notes

### Competing Interest Statement

The authors have declared no competing interest.

### Funding Statement

This study was funded by NIH/NIGMS (R01-GM124109) and NIH/NICHD P50 HD106446-01S1.

### Author Declarations

Ethics committee/IRB of Vanderbilt University Medical Center gave ethical approval for this work

