## Supplemental Table 1 for "Developing a natural language processing system using transformer-based models for adverse drug event detection in electronic health records"

**Supplemental Material**

**Supplemental Table 1.** Effects of Split Level on Model Performance using the n2c2 Dataset

| Model | Chunk Size | Split Level | Average of Precision | Average of Recall | Average of F-score |
| --- | --- | --- | --- | --- | --- |
| Bert-base-uncased | 10 chunks | Word | 0.721 | 0.714 | 0.716 |
|  |  | Sentence | 0.752 | 0.699 | 0.715 |
|  | 20 chunks | Word | 0.757 | 0.698 | 0.721 |
|  |  | Sentence | 0.806 | 0.730 | 0.761 |
| PubMedBERT | 10 chunks | Word | 0.758 | 0.710 | 0.728 |
|  |  | Sentence | 0.815 | 0.778 | 0.794 |
|  | 20 chunks | Word | 0.806 | 0.719 | 0.751 |
|  |  | Sentence | 0.838 | 0.804 | 0.823 |
| Clinical-Longformer | 10 chunks | Word | 0.808 | 0.772 | 0.787 |
|  |  | Sentence | 0.865 | 0.872 | 0.868 |
|  | 20 chunks | Word | 0.810 | 0.751 | 0.775 |
|  |  | Sentence | 0.850 | 0.850 | 0.850 |

The n2c2 dataset was processed with split-based approach at word and sentence levels. Across all models and chunk sizes, sentence level split performed consistently better than word level split, indicating that sentence level split keeping more complete context had a significant impact on improving model performance. Therefore, all results we reported using split-based approach were based on sentence-level splits. Default settings worked for most cases, and only in some cases where the model failed to converge, we made some adjustments to the batch size due to computational resource constraints. The performance of each model was evaluated by the average of three trials with different train-test split seeds for validation.
